# Quantitative Mass Spectrometry Analysis of Cerebrospinal Fluid Protein Biomarkers in Alzheimer’s Disease

**DOI:** 10.1101/2022.08.30.22279370

**Authors:** Caroline M. Watson, Eric B. Dammer, Lingyan Ping, Duc M. Duong, Erica Modeste, E. Kathleen Carter, Erik C. B. Johnson, Allan I. Levey, James J. Lah, Blaine R. Roberts, Nicholas T. Seyfried

## Abstract

Alzheimer’s disease (AD) is the most common form of dementia, with cerebrospinal fluid (CSF) β-amyloid (Aβ), total Tau, and phosphorylated Tau providing the most sensitive and specific biomarkers for diagnosis. However, these diagnostic biomarkers do not reflect the complex changes in AD brain beyond amyloid (A) and Tau (T) pathologies. Here, we report a selected reaction monitoring mass spectrometry (SRM-MS) method with isotopically labeled standards for relative protein quantification in CSF. Biomarker positive (AT+) and negative (AT-) CSF pools were used as quality controls (QCs) to assess assay precision. We detected 62 peptides (51 proteins) with an average CV of ∼13% across 30 QCs and 133 controls (cognitively normal, AT), 127 asymptomatic (cognitively normal, AT+) and 130 symptomatic AD (cognitively impaired, AT+). Proteins that could distinguish AT+ from AT-individuals included SMOC1, GDA, 14-3-3 proteins, and those involved in glycolysis. Proteins that could distinguish cognitive impairment were mainly neuronal proteins (VGF, NPTX2, NPTXR, and SCG2). This demonstrates the utility of SRM-MS to quantify CSF protein biomarkers across stages of AD.

## Background & Summary

Alzheimer’s disease (AD) affects more than 45 million people worldwide, making it the most common neurodegenerative disease^1^ (https://www.alz.org/media/Documents/alzheimers-facts-and-figures-2019-r.pdf; https://www.alzint.org/u/WorldAlzheimerReport2015.pdf). AD biomarker research has predominately focused on β-amyloid (Aβ) and Tau, as these proteins reflect pathological Aβ plaques and tau neurofibrillary tangles (NFT), respectively, in AD^2,3^. Although Aβ and Tau are the most sensitive and specific CSF biomarkers for diagnosis^4,5^ these two proteins do not reflect the heterogenous and complex changes in AD brain^6,7^. Furthermore, failed clinical trials of Aβ-based therapeutic approaches highlight the complexity of AD and the need for additional biomarkers to fully illustrate pathophysiology for advancements in diagnostic profiling, disease monitoring, and treatments^1,7^ (https://www.alz.org/media/Documents/alzheimers-facts-and-figures-2019-r.pdf; https://www.alzint.org/u/WorldAlzheimerReport2015.pdf).

Considering the diagnostic challenges related to the overlapping pathologies of neurodegenerative diseases, AD biomarkers that represent diverse pathophysiological changes could facilitate an early diagnosis, predict disease progression, and enhance the understanding of neuropathological changes in AD^1^. AD has a characteristic pre-clinical or asymptomatic period (AsymAD) where individuals have AD neuropathology in the absence of clinical cognitive decline^3,8,9^. Thus, biomarkers for the prodromal phase of AD that can begin changing years or decades before signs of cognitive impairment, would be valuable for disease intervention, clinical trial stratification, and monitoring drug efficacy.

Proteins are the proximate mediators of disease, integrating the effects of genetic, epigenetic, and environmental factors^7,10^. Network proteomic analysis has emerged as a valuable tool for organizing complex unbiased proteomic data into groups or “modules” of co-expressed proteins that reflect various biological functions, i.e., systems biology ^11-14^. The direct proximity of CSF to the brain presents a strong rationale to integrate the brain and CSF proteomes to increase the pathophysiological diversity among biofluid biomarkers of AD^5,15^. We recently integrated a human AD brain proteomic network with a CSF proteome differential expression analysis and revealed approximately 70% of the CSF proteome overlapped with the brain proteome^16^. Nearly 300 CSF proteins were identified as significantly altered between control and AD samples, representing predominately neuronal, glial, vasculature and metabolic pathways, creating an excellent list of candidates for further quantification and validation.

Here, we developed a high-throughput targeted selected reaction monitoring-based mass spectrometry (SRM-MS) assay^17^ to quantify and validate reliably detected CSF proteins in healthy individuals and individuals with asymptomatic or symptomatic AD for staging AD progression. We evaluated 200+ tryptic peptides that were selected using a data-driven approach from the integrated brain-CSF proteome network analysis. We selected peptides with differential abundance in AD CSF observed in >50 percent of case samples by discovery proteomics^16^ for synthesis as crude heavy standards. We used two pooled CSF reference standards to determine which peptides were reliably detected in CSF matrix. We reproducibly detected and reliably quantified 62 tryptic peptides from 51 proteins in 390 clinical samples and 30 pooled reference standards. Furthermore, using a combination of differential expression and receiver operating curve (ROC) analyses we found CSF proteins that can best discriminate stages of AD progression. Collectively, these data highlight the utility of a high throughput SRM-MS approach to quantify biomarkers associated with AD that ultimately hold promise for monitoring disease progression, stratifying patients for clinical trials, and measuring therapeutic response. Future studies will be necessary to assess the diagnostic and predictive utility of our CSF peptide SRM panel against gold-standard CSF (amyloid, tau and pTau) and imaging AD biomarkers in larger prospective patient cohorts.

## Methods

### Reagents and materials

Heavy labeled peptides (Thermo PEPotec SRM Peptide Libraries; Grade 2; crude as synthesized), trypsin, mass spectrometry grade, trifluoroacetic acid (TFA), foil heat seals (AB-0757), and low-profile square storage plates (AB-1127) were purchased from ThermoFisher Scientific (Waltham, MA). Lysyl endopeptidase (Lys-C), mass spectrometry grade was bought from Wako (Japan); sodium deoxycholate, CAA (chloroacetamide), TCEP (tris-2(-carboxyethyl)-phosphine), and triethylammonium hydrogen carbonate buffer (TEAB) (1 M, pH 8.5) were obtained from Sigma (St. Louis, MO). Formic acid (FA), 0.1% FA in acetonitrile, 0.1% FA in water, methanol, and sample preparation V-bottom plates (Greiner Bio-One 96-well Polypropylene Microplates; 651261) are from Fisher Scientific (Pittsburgh, PA). Oasis PRiME HLB 96-well, 30mg sorbent per well, solid phase extraction (SPE) cleanup plates were from Waters Corporation (Milford, MA).

### Pooled CSF as quality controls

Two pools of CSF were generated based on Aβ(1-42), total Tau, and pTau181 levels to create AD-positive (AT+) and AD-negative (AT-) quality control standards. Each pool consisted of approximately 50 mL of CSF by combining equal volumes of CSF selected from well characterized samples (∼45 unique individuals per pool) from the Emory Goizueta Alzheimer’s Disease Research Center (GADRC) and Emory Healthy Brain Study (EHBS). All research participants provided informed consent under protocols approved by the Institutional Review Board (IRB) at Emory University. CSF was collected by lumbar puncture and banked according to 2014 ADC/NIA best practices guidelines (https://www.alz.washington.edu/BiospecimenTaskForce.html). AD biomarker status for individual cases was determined on the Roche Elecsys® immunoassay platform^18-20^; the average CSF biomarker value is reported in parentheses. The control CSF pool (AT-) was comprised of cases with relatively high levels of Aβ(1-42) (1457.3 pg/mL) and low total Tau (172.0 pg/mL) and pTau181 (15.1 pg/mL). In contrast, the AD pool (AT+) was comprised of cases with low levels of Aβ(1-42) (482.6 pg/mL) and high total Tau (341.3 pg/mL) and pTau181 (33.1 pg/mL). The quality control (QC) pools were processed and analyzed identically to the CSF clinical samples reported.

### Clinical characteristics of the cohort

Human cerebrospinal fluid (CSF) samples from 390 individuals including 133 healthy controls, 130 patients with symptomatic AD, and 127 asymptomatic AD patients (cognitively normal but AD biomarker positive) were obtained from Emory’s GADRC and EHBS (**Fig. 1 and Table 1**). All symptomatic individuals were diagnosed by expert clinicians in the ADRC and Emory Cognitive Neurology Program, who are subspecialty trained in Cognitive and Behavioral Neurology, following extensive clinical evaluations including detailed cognitive testing, neuroimaging, and laboratory studies. CSF samples were selected to balance for age and sex (**Table 1**). For biomarker measurements, CSF samples from all individuals were assayed for Aβ(1-42), total Tau, and pTau using the Roche Diagnostics Elecsys® immunoassay platform^18-20^. The cohort characteristics are summarized in **Fig. 1 and Table 1**. Compared to our previous CSF studies^14,16,21^, there is minimal overlap with 329 of the 390 CSF samples (∼84%) unique to this study. Samples were stratified into controls, AsymAD, and AD based on Tau and Amyloid biomarkers status and cognitive score from the Montreal Cognitive Assessment (MoCA). All case metadata including disease state, age, sex, race, apolipoprotein (ApoE) genotype, MoCA scores, and biomarkers measurements were deposited on Synapse^22^.

**Figure 1.**
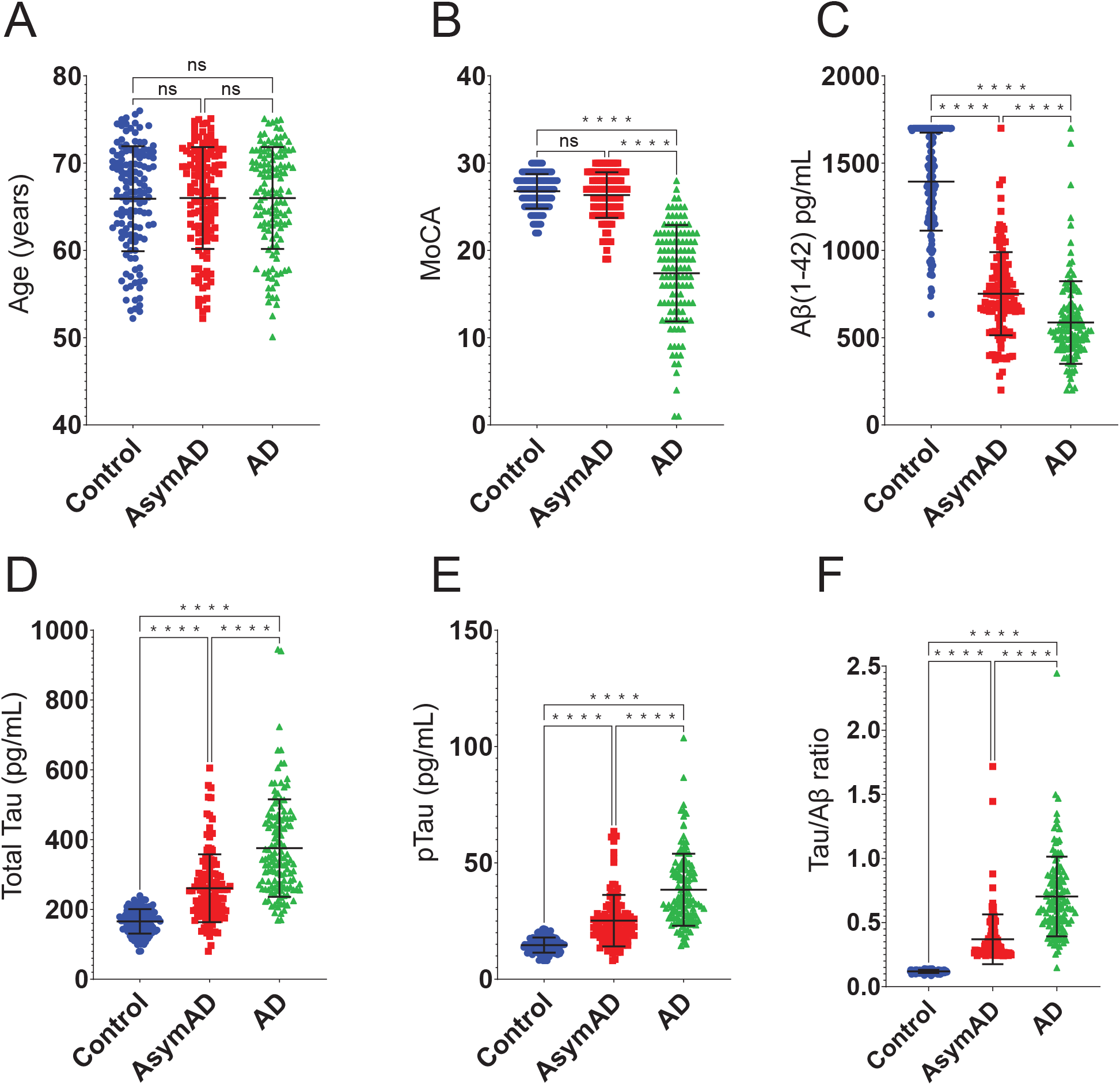
Cohort characteristics. A total of 390 samples (133 controls, 127 AsymAD, 130 AD unless otherwise noted) were analyzed using the following characteristics for grouping. (**A**) Age range across each group of the cohort was carefully selected to balance for age and sex (**Supplemental Table 1**). (**B**) Cognition was assessed using the Montreal Cognitive Assessment (MoCA) score; there is no significant difference in scores between the Control and AsymAD groups serving as the two cognitively normal diagnostic groups (133 controls, 127 AsymAD, 124 AD). The Roche Diagnostics Elecsys® platform was used for CSF biomarker measurements for Aβ(1-42) (**C**), Total Tau (133 controls, 127 AsymAD, 129 AD) (**D**), and pTau (**E**) (pg/mL) showing the significance between groups for each measurement. (**F**) Tau/Aβ ratio data across control, AsymAD and AD groups. There was no significant difference between AsymAD and AD groups to serve as our biomarker positive groups (133 controls, 127 AsymAD, 129 AD). The significance of the pairwise comparisons is indicated by overlain annotation of ‘ns’ (not significant; p>0.05) or asterisks; ****p≤0.0001.

**Table 1.** Cohort Characteristics

| Sample Group<br>Sample Size | CT<br>N=133 | AsymAD<br>N=127 | AD<br>N=130 |
| --- | --- | --- | --- |
| **Characteristics** |  |  |  |
| Sex | 99 F, 34 M | 94 F, 33 M | 97 F, 33 M |
| Age <sup>a</sup> | 66 ± 6 | 66 ± 6 | 66 ± 6 |
| MoCA <sup>b</sup> | 26.8 ± 2.0 | 26.4 ± 2.6 | 17.4 ± 5.5 |
| Aβ<sub>42</sub> <sup>b</sup> | 1394.0 ± 280.7 | 752.0 ± 238.2 | 587.0 ± 236.8 |
| tTau <sup>b</sup> | 165.8 ± 35.0 | 260.8 ± 97.2 | 375.7 ± 139.6 |
| pTau <sup>b</sup> | 14.7 ± 3.2 | 25.2 ± 11.1 | 38.5 ± 15.5 |

<sup>a</sup> Age in years. Values given as average ± standard deviation
<sup>b</sup> MoCA, Aβ<sub>42</sub>, tTau, and pTau in pg/mL. Values given in average ± standard deviation
Abbreviations: CT, Control; AD, Alzheimer's disease; MoCA, Montreal Cognitive Assessment

### Peptide selection and selected reaction monitoring assay

We harnessed both deep discovery and single-shot tandem mass tag (ssTMT) peptide data from CSF proteomics^14,16^. Here, we prioritized peptides for SRM validation that i) had one or more spectral match, ii) were differentially abundant (AD versus control) iii) or that mapped to proteins within brain-based biological panels that differed in AD^16^. Ultimately, we nominated 200+ peptides for synthesis as crude heavy standards. The heavy peptides contained isotopically labeled C-terminal lysine or arginine residues (^13^C, ^15^N) for each tryptic peptide. Based on the crude heavy peptide signal, the peptides were pooled to achieve total area signals ≥ 1×10^5^ in CSF matrix. The transition lists were created in Skyline-daily software (version 21.2.1.455)^23,24^. An in-house spectral library was created in Skyline based on tandem mass spectra from CSF samples. Skyline parameters were specified as: trypsin enzyme, Swiss-Prot background proteome, and carbamidomethylation of cysteine residues (+57.02146 Da) as fixed modifications. Isotope modifications included: ^13^C_6_^15^N_4_ (C-term R) and ^13^C_6_^15^N_2_ (C-term K). The top ten fragment ions that matched the criteria (precursor charges: 2; ion charges 1, 2; ion types: y, b) were selected for scrutiny. The top 5-7 transitions per heavy precursor were selected by manual inspection of the data in Skyline and scheduled transition lists were created for collision energy optimization. Collision energies were optimized for each transition; the collision energy was ramped around the predicted value in 3 steps on both sides, in 2V increments^25^. The selected transitions were tested in real matrix spiked with the heavy peptide mixtures. The three best transitions per precursor were selected by manual inspection of the data in Skyline and one scheduled transition list was created for the final assays. A list of transitions used in this study is deposited on Synapse^22^.

### Preparation of CSF for mass spectrometric analysis

All CSF samples were blinded and randomized. Each CSF sample was thawed and aliquoted into sample preparation V-bottom plates that also included quality controls. Each sample and quality control were processed independently in parallel. Crude CSF (50 μL) was reduced, alkylated, and denatured with tris-2(-carboxyethyl)-phosphine (5 mM), chloroacetamide (40 mM), and sodium deoxycholate (1%) in triethylammonium bicarbonate buffer (100 mM) in a final volume of 150 μL. Sample plates were heated at 95°C for 10 min, followed by a 10-min cool down at room temperature while shaking on an orbital shaker (300 rpm)^26^. CSF proteins were digested with Lys-C (Wako; 0.5 μg; 1:100 enzyme to CSF volume) and trypsin (Pierce; 5 μg; 1:10 enzyme to CSF volume) overnight in a 37°C oven. After digestion, heavy labeled standards for relative quantification (15 μL per 50 μL CSF) were added to the peptide solutions followed by acidification to a final concentration of 0.1% TFA and 1% FA (pH ≤ 2). Sample plates were placed on an orbital shaker (300 rpm) for at least 10 minutes to ensure proper mixing. Plates were centrifuged (4680 rpm) for 30 minutes to pellet the precipitated surfactant. Peptides were desalted with Oasis PRiME HLB 96-well, 30mg sorbent per well, solid phase extraction (SPE) cleanup plates from Waters Corporation (Milford, MA) using a positive pressure system. Each SPE well was conditioned (500 μL methanol) and equilibrated twice (500 μL 0.1% TFA) before 500 μL 0.1% TFA and supernatant were added. Each well was washed twice (500 μL 0.1% TFA) and eluted twice (100 μL 50% acetonitrile/0.1% formic acid). All eluates were dried under centrifugal vacuum and reconstituted in 50 μL mobile phase A (0.1% FA in water) containing Promega 6 × 5 LC-MS/MS Peptide Reference Mix (50 fmol/μL; Promega V7491).

### Liquid chromatography-tandem mass spectrometry (LC-MS/MS)

Peptides were analyzed using a TSQ Altis Triple Quadrupole mass spectrometer (Thermo Fisher Scientific). Each sample was injected (20 μL) using a 1290 Infinity II system (Agilent) and separated on an AdvanceBio Peptide Map Guard column (2.1×5mm, 2.7 μm, Agilent) connected to AdvanceBio Peptide Mapping analytical column (2.1×150mm, 2.7 μm, Agilent). Sample elution was performed over a 14-min gradient using mobile phase A (MPA; 0.1% FA in water) and mobile phase B (MPB; 0.1% FA in acetonitrile) with flow rate at 0.4 mL/min. The gradient was from 2% to 24% MPB over 12.1 minutes, then from 24% to 80% over 0.2 min and held at 80% B for 0.7 min. The mass spectrometer was set to acquire data in positive-ion mode using selected reaction monitoring (SRM) acquisition. Positive ion spray voltage was set to 3500 V for the Heated ESI source. The ion transfer tube and vaporizer temperatures were set to 325°C and 375°C, respectively. SRM transitions were acquired at Q1 resolution 0.7 FWHM, Q2 resolution 1.2 FWHM, CID gas 1.5 mTorr, 0.8 s cycle time.

### Data analysis

Raw files from Altis TSQ were uploaded to Skyline-daily software (version 21.2.1.455), which was used for peak integration and quantification by peptide ratios. QC SRM data were manually evaluated in Skyline by assessing retention time reproducibility, matching light and heavy transitions using Ratio Dot Product, and determining the peptide ratio precision using CV by QC condition. If Skyline could not automatically pick a consistent peak due to interference in the light transitions the peptide was removed from the analysis. Transition profiles were checked to insure the heavy and light transition profiles matched using the Ratio Dot Product value in Skyline. The Ratio Dot Product (1 = exact match) is a measure of whether the transition peak areas in the two label types are in the same ratio to each other. The average Ratio Dot Product value for each peptide was >0.90 for each QCs. If the retention time or Ratio Dot Product were outside of the expected range for a peptide in a few samples, the peaks were checked individually and adjusted as necessary. Total area ratios for each peptide were calculated in Skyline by summing the area for each light (3) and heavy (3) transition and dividing the light total area by the heavy total area. The Total Area Ratio CV was assessed using Skyline and the peptide was removed from the analysis if the CV>20% by QC condition. Next, the individual CSF samples were analyzed in a blinded fashion. We used the total area ratios (peptide ratios) for each targeted peptide in each sample and QC analysis; therefore, there are no missing values in the Data Matrix on Synapse. The raw data files and Skyline file have been deposited on Synapse^22^.

### Statistical analyses

We used Skyline-daily software (version 21.2.1.455) and GraphPad Prism (version 9.4.1) software to calculate means, medians, standard deviations, and coefficients of variations^24^. Peptide abundance ratios were log2-transformed, and zero values were imputed as one-half the minimum nonzero abundance measurement. Then, one-way ANOVA with Tukey *post hoc* tests for significance of the paired groupwise differences across diagnosis groups was performed in R (version 4.0.2) using a custom calculation and volcano plotting framework implemented and available as an open-source set of R functions documented further on https://www.github.com/edammer/parANOVA. T test p values and Benjamini-Hochberg FDR for these are reported for two total group comparisons, as was the case for AT+ versus AT-peptide mean difference significance calculations. Receiver-operating characteristic (ROC) analysis was performed in R (version 4.0.2) with a generalized linear model binomial fit of each set of peptide ratio measurements to the binary case diagnosis subsets AD/Control, AsymAD/Control, and AD/AsymAD using the pROC package implementing ROC curve plots, and calculations of AUC and AUC DeLong 95% confidence interval. Additional ROC curve characteristics including sensitivity, specificity, and accuracy were calculated with the reportROC R package. Robustness of the ROC calculations of AUC were confirmed using k-fold cross validation (k=10 folds, with each fold containing case subsets with equal distributions of the binary outcome) implemented using the cvAUC R package functions for calculating cross-validated AUC (cvAUC), and confidence interval on pooled predictions, and these calculations were consistently within 1 percent of AUC as calculated using a single calculation on the full data (data not shown). Venn diagrams were generated using the R vennEuler package, and the heatmap was produced using the R pheatmap package/function. R boxplot function output was overlaid with beeswarm-positioned individual measurement points using the R beeswarm package. Pearson correlations of SRM peptide measurements to immunoassay measurements of Aβ(1-42), total Tau, phospho-T181 Tau, and the ratio of total Tau/ Aβ were performed using the corAndPvalue WGCNA function in R. Correlation scatterplots were generated using the verboseScatterplot WGCNA function.

## Data Records

All files have been deposited on Synapse^22^. The data folder contains sample traits, transition details, the peptide ratio data matrix, transition ratios, and protein details. The Metadata file contains the available traits for each sample analyzed including: SampleID – Internal Sample Identifier; Age(years) – Deidentified Age in years; Sex – Binary Sex; Race – self identified race; Educ – formal years of education; MoCA – Montreal Cognitive Assessment Score ranging from 0-30; APOE status – APOE genotype; Aβ42, tTau, pTau – as measured in CSF by Roche Elecsys® immunoassay platform; tTau:Aβ42 – ratio of tTau/ Aβ42; SampleRunOrder – the order the samples were acquired; Condition – the Control/AsymAD/AD group used in the analysis. The DataMatrix file contains peptide ratio data for each sample. Data file names are list in the top row of the DataMatrix: AT+ QC samples begin with AD_; AT-QC samples begin with CTL_; Individual samples can be mapped back to the Metadata using the sample run order (Sxxx) or SampleID (_xxxxx); Plate/box and well position also defined in the file name (_Bx_Axx). The MS RAW files folder contains all mass spectrometry raw files (N=423) from both quality control replicates and clinical samples. The Skyline quantification folder contains the Skyline file that was used to report the peptide ratios, calculate CVs and means, and ratio dot product.

## Technical Validation

### Assessing peptide precision using pooled CSF quality control (QC) standards

We generated two pools of CSF reference standards as QCs based on biomarker status (AT- and AT+). These QCs were processed and analyzed (at the beginning, end, and after every 20 samples per plate) identically to the individual clinical samples for testing assay reproducibility. We analyzed 30 QCs (15 AT- and 15 AT+) over approximately 5 days during the run of clinical samples. We identified 62 peptides from 51 proteins as reliably measured in the pooled reference standards. Notably, only 5 of these peptides overlap with previous published PRM dataset given the unique differences in sample preparation, MS platform and peptide selection^21^. We included 58 peptides from 51 proteins in our biomarker analysis, plus peptides specific for the four APOE alleles for proteogenomic confirmation of APOE genotypes^27,28^. The technical coefficient of variation (CV) of each peptide was calculated based on the peptide area ratio for the biomarker negative (AT-) and positive (AT+) QCs. We defined CSF peptide biomarkers with CVs ≤ 20% as quantified with high precision in these technical replicates which were un-depleted and unfractionated CSF sample pools. Technical and process reproducibility for all reported peptides was below 20% (CV < 20%) in at least one pooled reference standard. The average CVs for all peptides in the AT- and AT+ QCs were 13% and 12%, respectively. **Supplemental Table 1** contains the QC statistics for the biomarker and APOE allele specific peptides. Levels of HBA and HBB peptides can be used to assess the levels of potential blood contamination^29^ in each of the CSF samples across individual plates (**Supplemental Fig. 1**). Correction for blood contamination could improve the statistics; however, no correction was performed for the statistical analyses presented. We used the protein directions of change to assess accuracy in the QC pools. The volcano plot between peptides measured in the pools highlights peptide/protein levels that are consistent with previously reported AD biomarkers (**Supplemental Fig. 2**)^16,21^.

### Monitoring LC-MS/MS instrument performance

The sample reconstitution solution contained Promega 6×5 LC-MS/MS Peptide Reference Mix (50 fmol/μL)^30^. The Promega Peptide Reference Mix provides a convenient way to assess LC column performance and MS instrument parameters, including sensitivity and dynamic range. The mix consists of 30 peptides; 6 sets of 5 isotopologues of the same peptide sequence, differing only in the number of stable, heavy-labeled amino acids incorporated into the sequence using uniform 13C and 15N atoms making them chromatographically indistinguishable. The isotopologues were specifically synthesized to cover a wide range of hydrophobicities so that dynamic range could be assess across the gradient profile (**Fig. 2A**). Each isotopologue represents a series of 10-fold dilutions, estimated to be 1 pmole, 100 fmole, 10 fmole, 1 fmole, and 100 amole for each peptide sequence in a 20 μL injection, a range that would challenge the lowest limits of detection of the method (**Fig. 2B**). We assessed the raw peak areas in 423 injections over 5 days to determine the label-free CV for each peptide isotopologue (**Fig. 2B**). The 100-amole level (0.0001x) was not detected (ND) for any of the peptide sequences. Based on the label-free CV, we determined the lowest limit of detection for each peptide to be between 1-10 fmole across the gradient profile with a dynamic range spanning 4 orders of magnitude for all peptides except the latest eluting peptide at 13.3 minutes (**Fig. 2C**).

**Figure 2.**
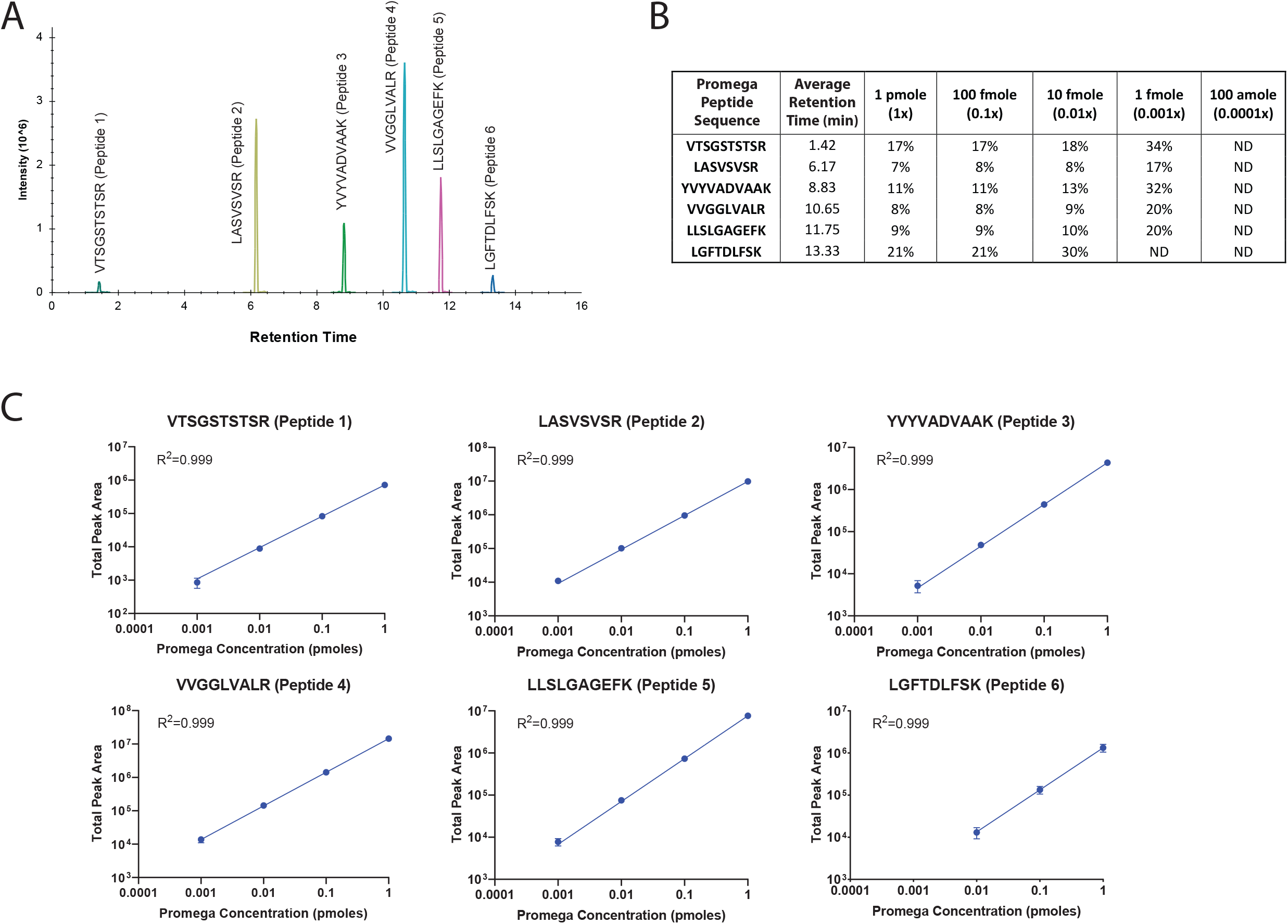
Isotopologue peptide internal reference standards to determine consistency of LC-MS/MS platform. Each of the CSF samples were spiked with a six-peptide, 5 isotopologue concentration LC-MS/MS Peptide Reference Mix from Promega (50 fmol/μL). (**A**) Extracted ion chromatogram for the 6 peptide (1pmol) mixture illustrating the wide range of retention times due to their hydrophobicity. (**B**) The raw peak areas in 423 injections over 5 days were used to determine the label-free CV for each peptide isotopologue estimating the lowest limits of detection to be between 1-10 fmole for each peptide. (**C**) The 5 unique isotopologues are used to assess the dynamic range across the gradient profile and each peptide demonstrates linearity across 3-4 orders of magnitude in the batch of 423 injections. Error bars represent the standard deviation across 423 injections.

### Technical replicate variance

Three individual samples were analyzed in duplicate scattered throughout the sample run sequence to assess technical replicate variance. We graphed the log2(ratio) for each of 58 biomarker peptides in replicate 1 versus replicate 2 for each sample and determined the Pearson correlation coefficient with associated P value (**Supplemental Fig. 3**). The analysis showed a near-identical correlation (ρ=0.996-0.998; p<1e-200) between each of the technical replicate pairs for the three individual CSF samples, supporting the same high level of method reproducibility we found using the QC pools.

### Concordance between a discovery (ssTMT) and replication (SRM) datasets

Since our peptide targets were largely based on multiple ssTMT datasets^16^, we generated a representative ssTMT peptide level volcano from one of these datasets comprised of 297 individuals (147 control and 150 AD) (**Fig. 3A**). There are 44 of 62 SRM peptides that overlap with this ssTMT dataset and are highlighted in yellow on the volcano plot (**Fig. 3A**). To establish peptide concordance, we also compared the direction of change or effect size (log2 fold change) for 40 overlapping peptides, excluding albumin, hemoglobin, and APOE allele specific peptides. **Fig. 3B** shows significant correlation (cor = 0.91; p = 2.8e-15) between SRM and ssTMT peptide highlighting the accuracy and concordance of measurements across both MS assays. Thus, despite substantial differences in chromatography (nanoflow versus standard flow), MS instrumentation (Orbitrap versus triple quadrupole), and protein quantitation approaches (ssTMT versus SRM), the selected peptides in this assay are highly reproducible and robust in their direction of change in AD CSF.

**Figure 3.**
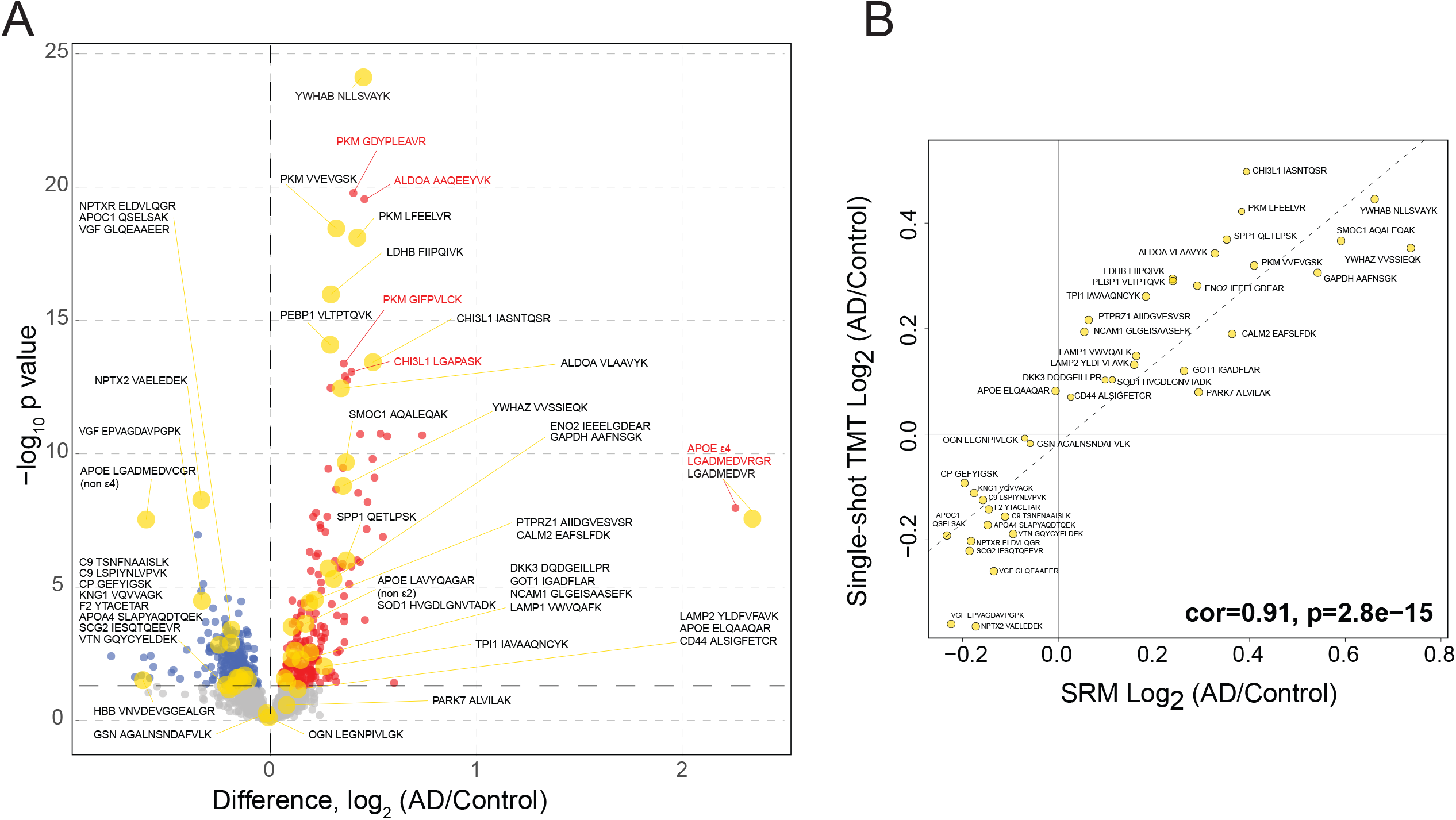
Peptide concordance between SRM and ssTMT datasets.0. (**A**) Volcano plot displaying the log_2_ fold change (FC) (x-axis) against t-test log10 p-value (y-axis) for all peptides (n=2,340) comparing AD (n=150) versus Controls (n=147). Cutoffs were determined by significant differential expression (p<0.05) between control and AD cases. Peptides with significantly decreased levels in AD are shown in blue while peptides with significantly increased levels in disease were indicated in red. 44 of 62 SRM peptides that overlap with this ssTMT dataset and SRM are highlighted as larger yellow points with black text labels. Red text and traces to red points are labels for peptides not included in the current SRM study that were significantly upregulated in the ssTMT dataset. (**B**) Correlation between the fold-change (AD vs control) of all selected overlapping peptides (n=40) across SRM (x-axis) and ssTMT (y-axis) were strongly correlated (cor=0.91, p=2.8e−15).

## Usage Notes

This targeted mass spectrometry dataset serves as a valuable resource for a variety of research endeavors including, but not limited to, the following applications:

### Use case 1: Peptide abundance in CSF

This dataset provides a reference for peptide detectability in CSF under relatively high-throughput conditions, especially if an investigator wants to determine whether their protein of interest has abundance above the lower limit of detection in CSF under these analytical conditions. Raw data deposited on Synapse^22^ contains transitions for over 200 peptides that were robustly detected in CSF discovery proteomics^16,21^.

### Use case 2: Using APOE allele specific peptides for genotyping

Apolipoprotein E (ApoE) has three major genetic variants (E2, E3, and E4, encoded by the ε2, ε3 and ε4 alleles, respectively) that differ by single amino acid substitutions^31^. APOE genotype is closely related to AD risk^32^ with ApoE4 having the highest risk, ApoE2 the lowest risk, and ApoE3 with intermediate risk^33,34^. Due to the amino acid substitutions in each variant, there are allele specific peptides that can be targeted by mass spectrometry^27,35^. We monitored CLAVYQAGAR (APOE2), LGADMEDVR (APOE4), LGADMEDVCGR (APOE2 or APOE3), and LAVYQAGAR (APOE3 or APOE4) to confirm the APOE genotype of each CSF sample in a concurrent SRM-MS method^22^. The CV for each APOE peptide in each QC is listed in **Supplemental Table 1**. Previous studies report the association of APOE genotype with various clinical, neuroimaging, and biomarker measures^36-39^. Exploring the relationship between APOE status and the CSF biomarker peptides presented requires further analysis reserved for future studies.

### Use case 3: Stage-specific differences in peptide and protein levels

The described cohort includes control, AsymAD, and AD groups across the Amyloid/Tau/Neurodegeneration (AT/N) framework^40^, which allows for the comparison of peptide and protein differential abundance across stages of disease. Investigators can focus on comparisons that are specific to symptomatic AD or those with potential for staging AD by using the AsymAD group compared to the control group. By comparing candidate biomarkers using ANOVA (excluding APOE allele specific peptides), we found 41 differentially expressed peptides (36 proteins) in AsymAD vs controls (**Fig. 4A**), 35 differentially expressed peptides (30 proteins) in AD versus controls (**Fig. 4B**), and 21 differentially expressed peptides (18 proteins) in AD vs AsymAD (**Fig. 4C**). The Venn diagram summarizes the differentially expressed peptides across groups in **Fig. 4D**.

**Figure 4.**
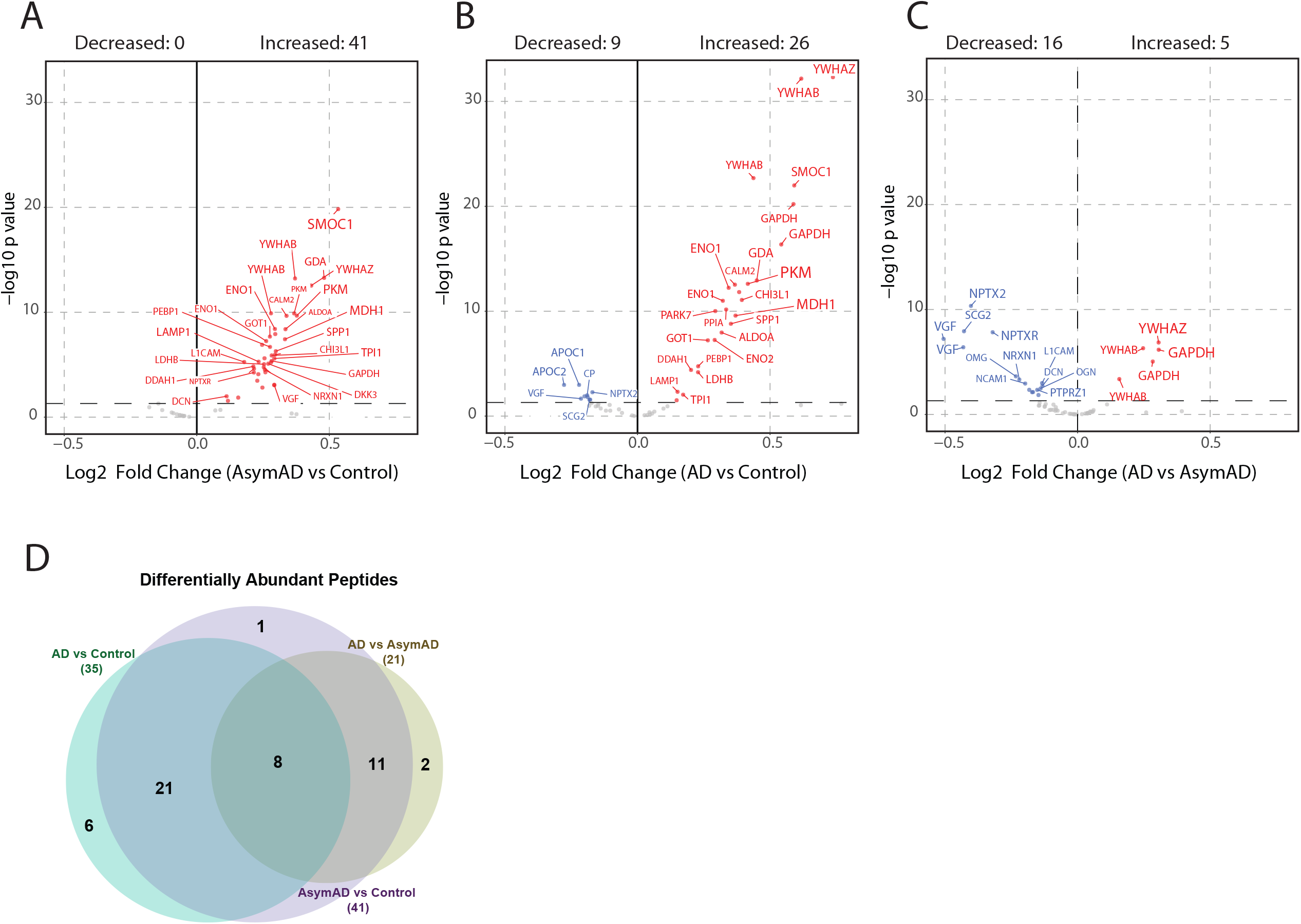
Differential expression analysis across stages of AD. ANOVA analysis with Tukey *post hoc* FDR was performed for pairwise comparison of mean log_2_(ratio) differences between the 3 stages of AD (i.e., Control, AsymAD and AD) of N=390 total case samples and plotted as a volcano. Significance threshold for counting of peptides was p < 0.05 (dashed horizontal line). Differentially expressed peptides for (**A**) AsymAD (N=127) versus control (N=133), (**B**) AD (N=130) versus control, and (**C**) AD versus AsymAD are labeled by their gene symbols. (**D**) Counts of peptides with significant difference in any of the 3 dichotomous comparisons are presented as a Venn diagram. Full statistics from the ANOVA and Tukey post-hoc analysis is presented in **Supplemental Table 2**.

Using a differential abundance analysis, we were able to stratify the changing proteins as early or progressive biomarkers of AD (**Fig. 4 and 5**). The log_2_-fold change (Log_2_ FC) from the volcano plots in **Fig. 4** are represented as a heatmap in **Fig. 5A** to illustrate how each peptide is changing across each group comparison. Twenty-two peptides (21 proteins) were early biomarkers of AD because they were significantly different in AsymAD versus controls, but not significantly different in AD versus AsymAD (**Fig. 5A**). A plurality of these proteins mapped to metabolic enzymes linked to glucose metabolism (PKM, MDH1, ENO1, ALDOA, ENO2, LDHB, and TPI1, also in **Supplemental Table 2**)^13,14^. SMOC1 and SPP1, markers linked to glial biology and inflammation^14,16^, were also increased in AsymAD samples compared to controls (**Fig. 5B**, *top row*). GAPDH, YWHAB and YWHAZ proteins were found to be progressive biomarkers of AD because the proteins were differentially expressed from Control to AsymAD and from AsymAD to AD with a consistent trend in direction of change (**Fig. 5B**, *middle row*). Proteins associated with neuronal/synaptic markers including VGF, NPTX2, NPTXR, and L1CAM were increased in AsymAD compared to controls but decreased in AD vs controls (**Fig. 5B**, *lower row*). Interestingly, we found 14 peptides (13 proteins) that were up in AsymAD as compared to Control but down in AD when compared to AsymAD. A majority of these proteins map to neuronal/synaptic markers including VGF, NPTX2, NPTXR, which are some of the most correlated proteins in post-mortem brain to an individual’s slope of cognitive trajectory in life (**Fig. 5A and 5B**, *lower row*)^41^.

**Figure 5.**
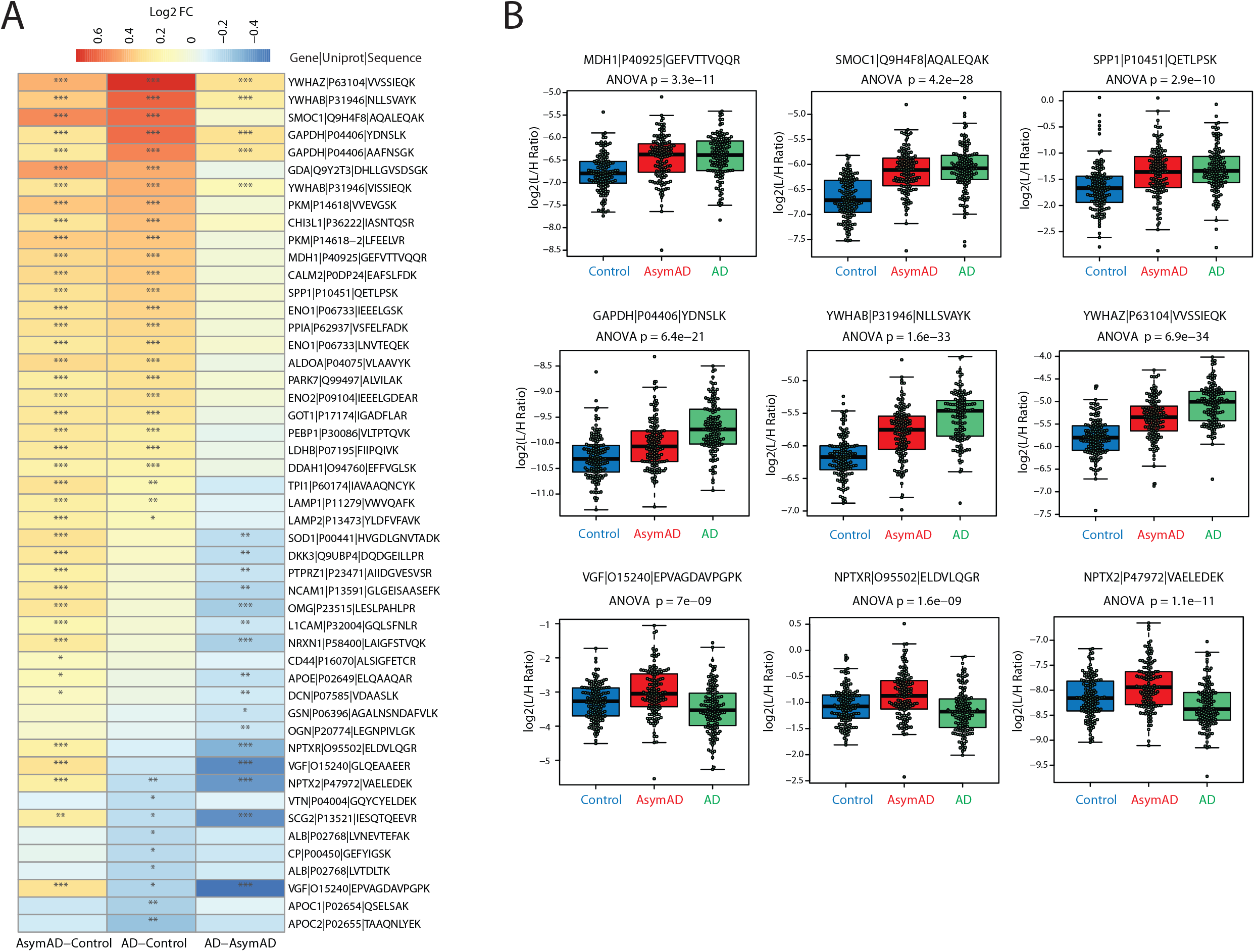
Stratifying early from progressive biomarkers of AD. (**A**) The magnitude of positive (red) and negative (blue) changes are shown on a gradient color scale heatmap representing mean log_2_-fold change (Log_2_FC) for each of 49 peptides significant in any of the 3 group comparisons. Tukey significance of the pairwise comparisons is indicated by overlain asterisks; *p<0.05, **p<0.01, ***p<0.001. (**B**) Peptide abundance levels of selected panel markers that are differentially expressed between groups. The upper row highlights biomarkers that are significantly different in AsymAD versus controls, but not significantly different in AsymAD versus AD. The middle row of 3 peptides highlights progressive biomarkers of AD, which show a stepwise increase in abundance from control to AsymAD to AD cases. The bottom row highlights a set of proteins that are increased in AsymAD compared to controls but decreased in AD versus control or AsymAD samples.

### Use case 4: Correlation of peptide biomarker abundance to Aβ(1-42), Tau, pTau and cognitive measures

The comparison of existing biomarkers to the SRM peptide measurements can be accomplished by correlation, where the degree of correlation indicates how similar a peptide measurement is to the established immunoassay-measured biomarkers of Aβ(1-42), total Tau, and pTau as well as cognition (MoCA score). In **Fig. 6A**, we demonstrate that 57 of the 58 biomarker peptides have significant correlation to at least one of the above biomarkers, or the ratio of total Tau/Aβ. Individual correlation scatterplots and linear fit lines for three of the peptides (SMOC1: AQALEQAK, YWHAZ: VVSSIEQK, and VGF: EPVAGDAVPGPK) are provided in **Fig. 6B**. Significant correlations of these peptides to the established biomarker and cognitive measures indicate the potential of these measurements to classify or stage disease progression. The targeted SRM measurement correlations largely agree with those observed from unbiased discovery proteomics^42^ and parallel reaction monitoring^21^ experiments.

**Figure 6.**
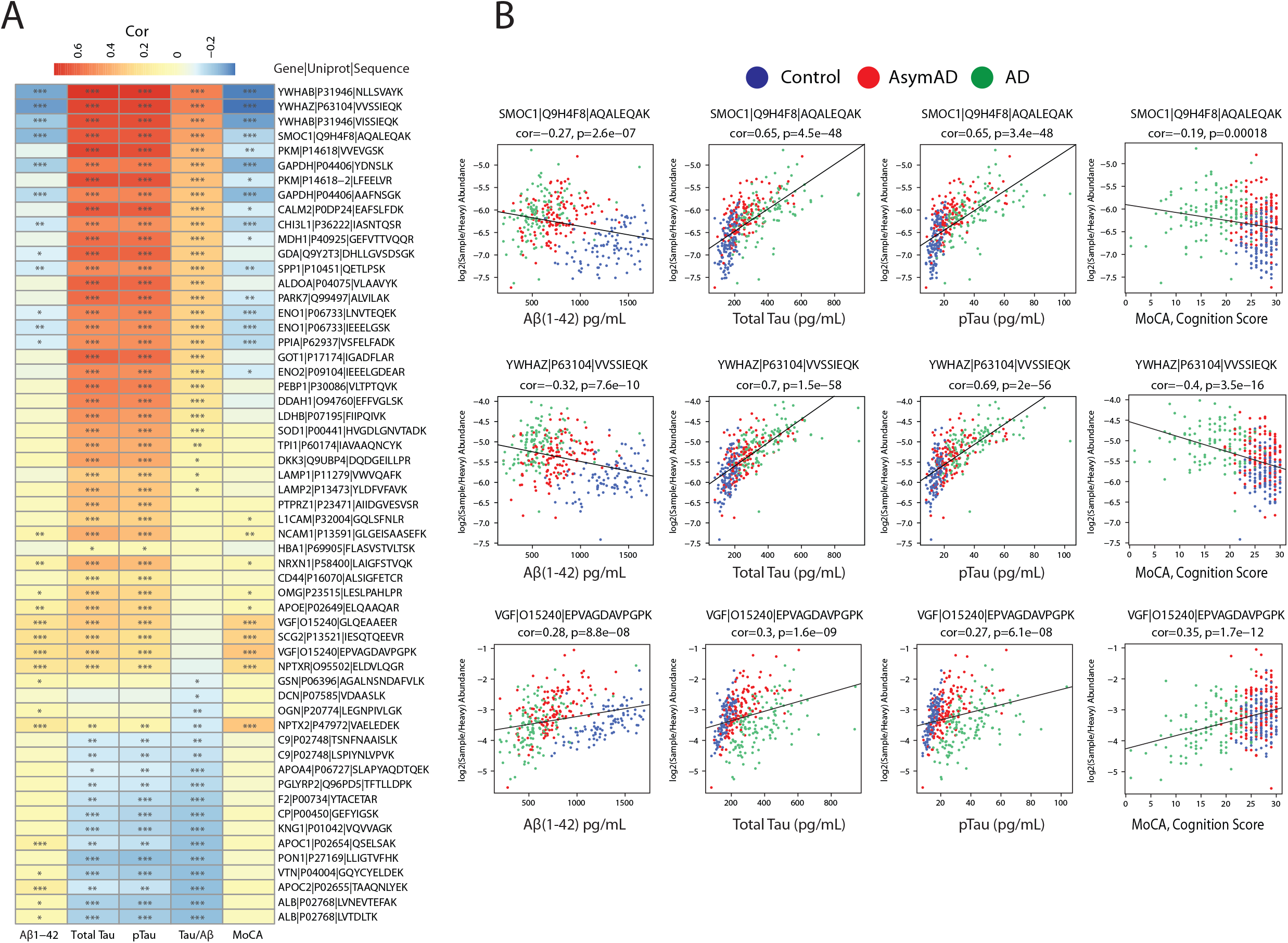
Correlating CSF peptide biomarker abundances to amyloid, Tau, and cognitive measures. (**A**) Positive (red) and negative (blue) Pearson correlations between biomarker peptide abundance and immunoassay measures of Aβ(1-42), total Tau, phospho-T181 Tau (pTau), ratio of total Tau/Aβ and cognition (MoCA score). Student’s *t* test significance is indicated by overlain asterisks; *p<0.05, **p<0.01, ***p<0.001. (**B**) Individual correlation scatterplots are shown for SMOC1 (upper row), YWHAZ (middle row), and VGF (lower row). Individual cases are colored by their diagnosis; blue for controls, red for AsymAD cases, and green for AD cases. Amyloid immunoassay measures of 1,700 (maximum, saturated value in the assay) were not considered for correlation.

### Use case 5: Receiver-operating characteristic (ROC) analysis for evaluating biomarker diagnostic capability

The capacity for peptide measurements to serve as a diagnostic biomarker distinguishing individuals with AD and even asymptomatic disease from individuals not on a trajectory to develop AD is well-established, with secreted amyloid and tau peptide measurements in CSF being the current gold standard for interrogation of patients’ AD stage from their CSF^43^ where CSF Aβ(1-42) concentration inversely correlates to plaque deposition in the living brain^44^. The measurements of additional peptides collected here are appropriate for comparison to immunoassay measurements of CSF amyloid and Tau biomarker positivity, or a dichotomized cognition rating, or other ancillary traits such as diagnosis for the 390 individuals. To demonstrate this utility, we performed receiver-operating characteristic (ROC) curve analysis and calculated the area under the curve (AUC) for all 62 peptide measures as fitting a logistic regression to 3 subsets of samples divided to represent known pairs of disease stages, namely AD versus control, AsymAD versus control, and AD vs AsymAD (**Fig. 7** and **Supplemental Table 4**). The top performing peptide for the YWHAZ gene product 14-3-3 ζ protein demonstrated an AUC of 89.5% discrimination of AD from control cases consistent with previous studies^21,45,46^. SMOC1 AUC of 81.8% was the best performing peptide for discrimination of AsymAD from control groups. In contrast, the synaptic peptides to NPTX2 (AUC of 74.0%), NPTXR (AUC of 71.1%), VGF (AUC of 70.1%) and SCG2 (AUC of 69.8%) best discriminated AD from AsymAD groups suggesting that neurodegeneration due to AD pathology is occurring in the symptomatic phase of disease^47^. **Fig. 7** shows the top five peptides by AUC for each of the three comparisons, highlighting the potential of this data set to aid in the design or validation of stage-specific biomarkers. Additional future analysis using these peptides alone or in combination could be used to subtype, predict disease onset, and gauge treatment efficacy.

**Figure 7.**
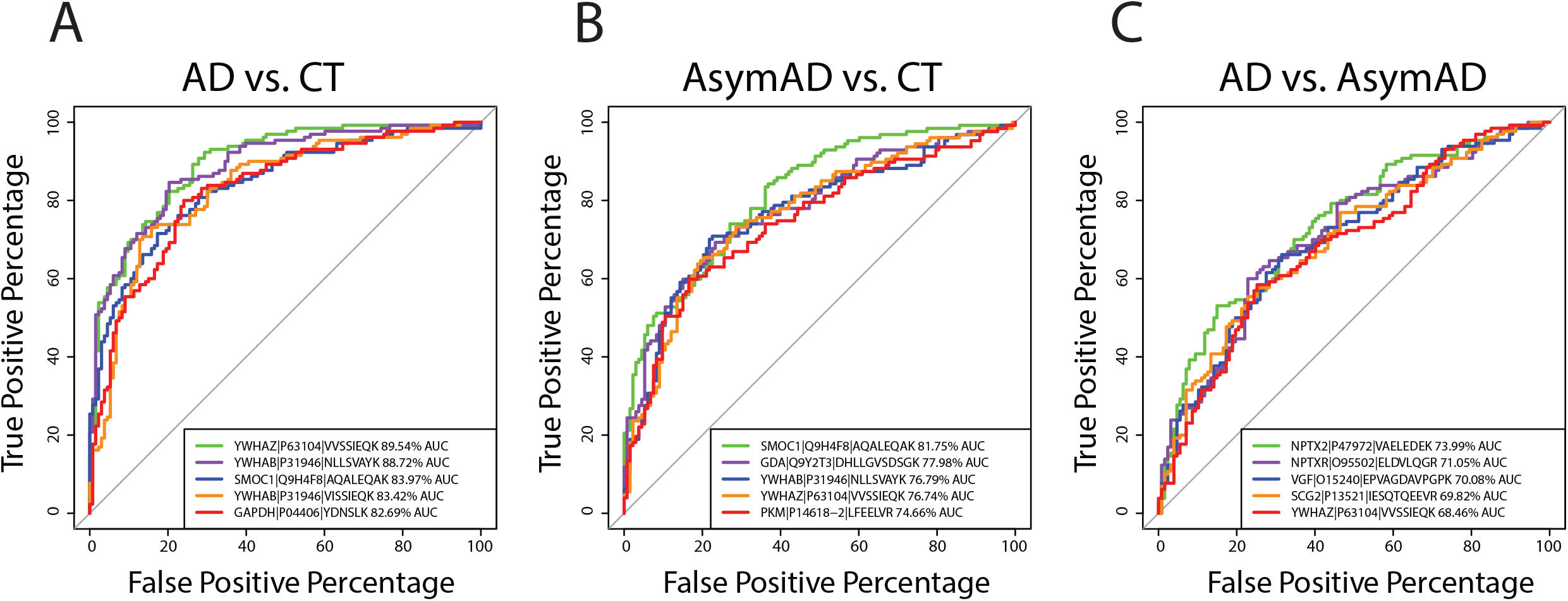
Receiver-operating characteristic (ROC) curve analysis of peptide diagnostic potential. ROC curves for each of three pairs of diagnosed case groups were generated to determine the top-ranked diagnostic biomarker peptides among the 58-peptide panel plus 4 APOE specific peptides. (**A**) A total of 263 AD (N=130) and control (N=133) CSF case samples were classified according to the logistic fit for each peptide’s log_2_(ratio) measurements across these samples, and the top 5 ranked by AUC are shown. (**B**) Top five performing peptides for discerning AsymAD (N=127) from control (N=133) case diagnosis groups are provided with AUCs, nominating these peptides as potential markers of pre-symptomatic disease, and as cognates for AT+ biomarker positivity. (**C**) Symptomatic AD (N=130) and AsymAD (N=127) discerning peptides were ranked by AUC and the top five ROC curves are shown and nominated as cognate CSF measures for compromised patient cognition.

## Data Availability

Raw mass spectrometry and pre- and post- processed peptide abundance data can be found at https://www.synapse.org/#!Synapse:syn34054965. The results published here are in whole or in part based on data obtained from the AMP-AD Knowledge Portal (https://adknowledgeportal.synapse.org) by the AMP-AD Target Discovery Program and other programs supported by the National Institute on Aging to enable open-science practices and accelerate translational learning.

## Code Availability

Custom code generating use case figures and tables including correlation plots, volcanoes, Venn diagram, annotated heatmap, statistics tables, and ROC curves is available for download with registration for a free account on synapse.org. The code is available as R scripts from https://doi.org/10.7303/syn35927166 (the Analysis folder) deposited on Synapse^22^. These scripts were run as provided on R version 4.0.2 with the two provided input files to generate outputs.

## Acknowledgements

This study was supported by the following National Institutes of Health funding mechanisms: U01AG061357 (A.I.L and N.T.S), R01AG070937-01 (J.J.L) and P30AG066511 (A.I.L). We acknowledge our colleagues at Emory for providing critical feedback.

## Author contributions

Conceptualization, C.M.W., L.P., D.M.D., J.J.L., A.I.L., B.R.R, N.T.S.; Methodology, C.M.W., E.B.D, L.P., D.M.D., E.M., E.K.C., B.R.R, N.T.S.; Investigation and Formal Analysis, C.M.W., E.B.D, L.P., D.M.D., B.R.R, N.T.S.; Writing – Original Draft, C.M.W., E.B.D., N.T.S.; Writing – Review and Editing, C.M.W., E.C.B.J, B.R.R., J.J.L., A.I.L., N.T.S.; Funding Acquisition, A.I.L. and N.T.S.; Supervision, N.T.S.

## Supplemental Data

**Supplemental Figure 1.**
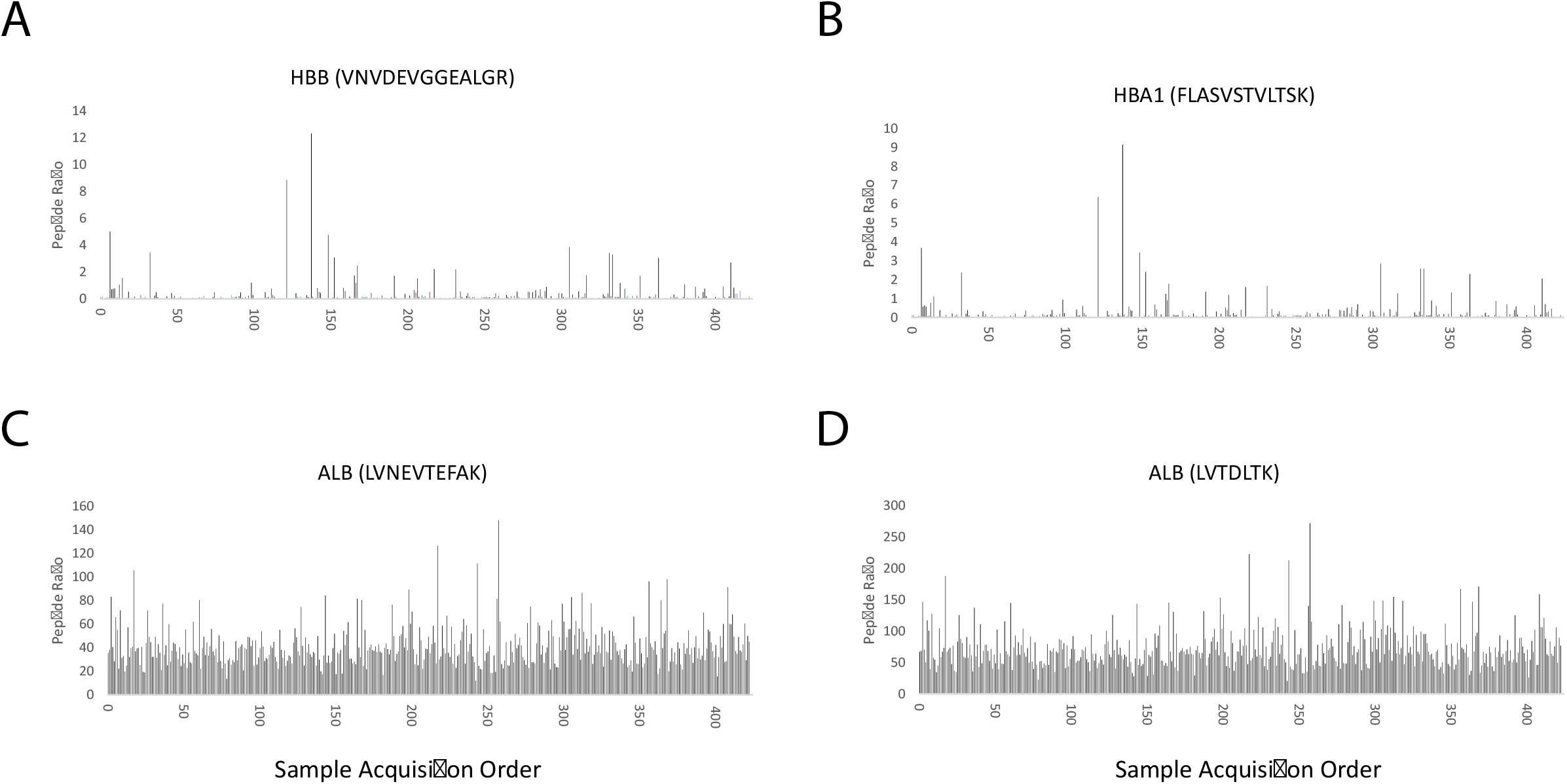
Monitoring background peptide levels in CSF. Three proteins were monitored for levels of potential blood contamination in each of the CSF samples. The peptide ratio for hemoglobin subunit alpha (**A**), hemoglobin subunit beta (**B**), and albumin (**C** and **D**) peptides are plotted for each of the CSF samples (N=423) in acquisition order.

**Supplemental Figure 2.**
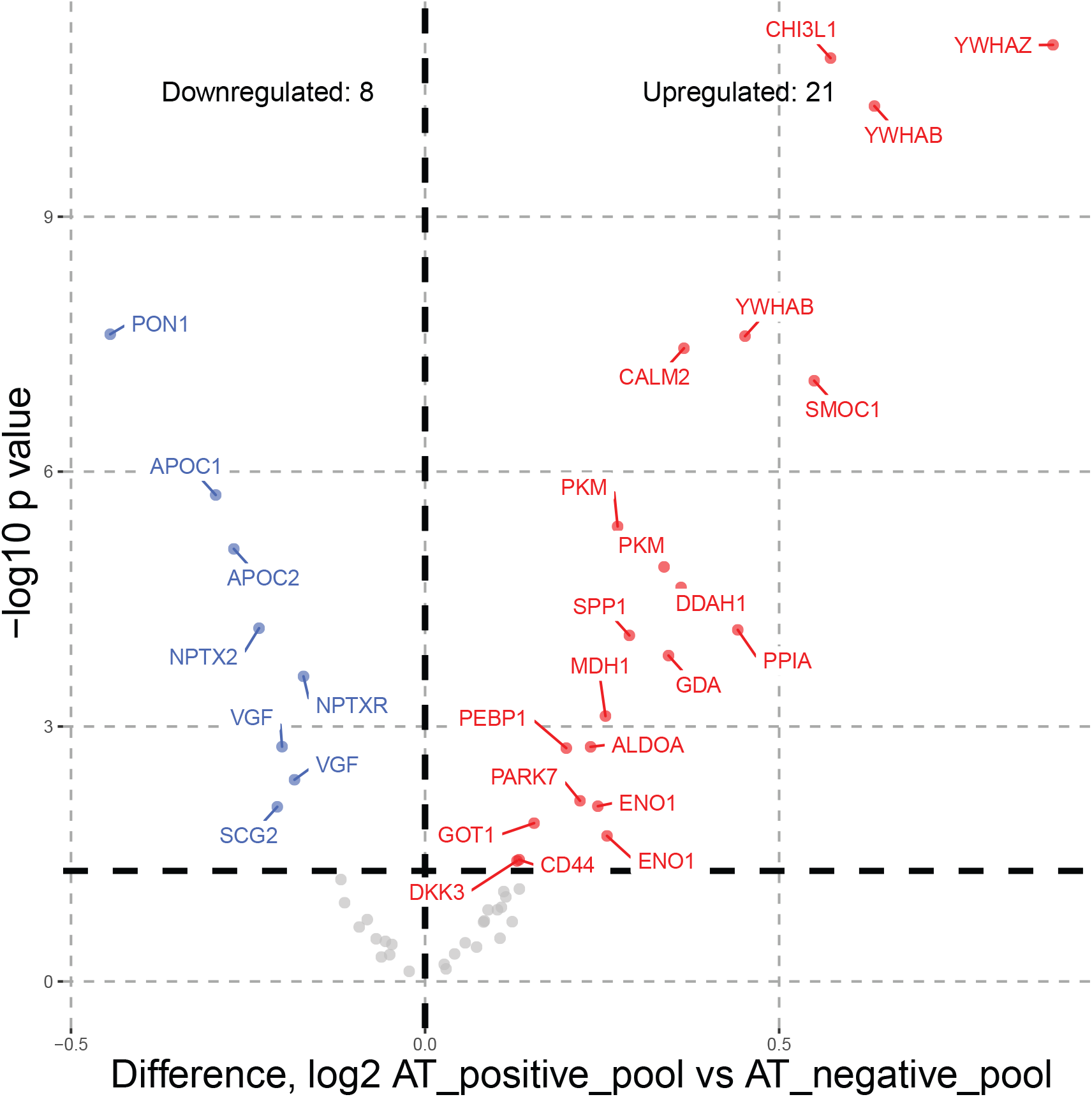
Differentially abundant peptides representing changed proteins in AT-vs AT+ QC CSF pools. The differentially abundant proteins in the QC pools were used to check the accuracy of the fold change consistent with our other studies^16^. We found 21 upregulated and 10 downregulated peptides. This result validated the direction of change of six proteins nominally significantly downregulated in previously published discovery proteomics (PON1, APOC1, NPTX2, VGF, NPTXR, and SCG2), and of sixteen proteins previously reported as upregulated (YWHAZ, GDA, CHI3L1, PKM, CALM2, SMOC1, YWHAB, MDH1, ALDOA, ENO1, GOT1, PPIA, DDAH1, PEBP1, PARK7, and SPP1)^16,21^.

**Supplemental Figure 3.**
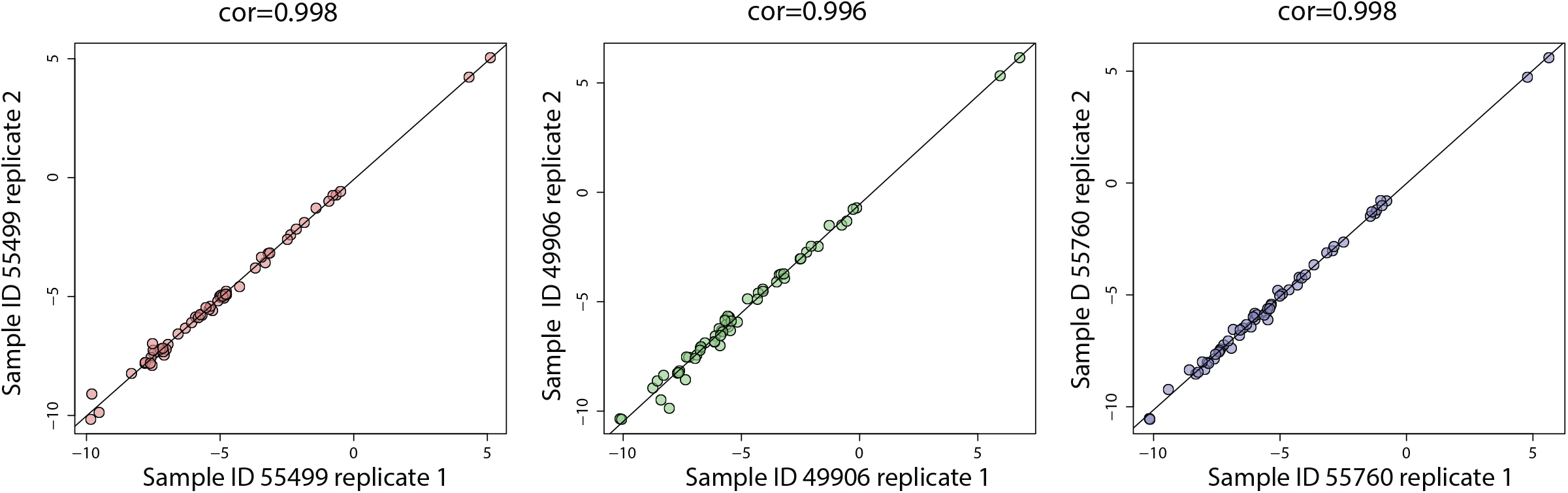
Technical reproducibility of peptide measurements in replicate CSF samples. Pearson correlation and p-value of replicate measures of 58 peptides in the 3 replicated CSF samples that were analyzed randomly within the series of 423 injections by SRM-MS.

**Supplemental Table 1**. Coefficient of variation (CV) values for 58 biomarker peptides and APOE allele specific peptides in AT- and AT+ QC pools.

**Supplemental Table 2**. ANOVA of differential abundance analysis for 58 biomarker peptides across Control, AsymAD and AD sample pairwise group comparisons. The SRM proteins were also cross-referenced with protein data and module membership from Johnson et al. Consensus Network^48^, and mapped to CSF protein panels from Higginbotham et al. Integrated Proteomic analysis^16^.

**Supplemental Table 3**. Pearson correlations (rho), Student p values of correlation significance, and numbers of paired observations for correlation of biomarker peptide abundances to immunoassay measures of Aβ(1-42), total Tau, phospho-T181 Tau, and the ratio of total Tau/ Aβ.

**Supplemental Table 4**. ROC curve statistics including AUC, p, 95% DeLong confidence interval, accuracy, specificity, and sensitivity for dichotomous diagnosis case sample groups.

## References

1 Scheltens, P. et al. Alzheimer’s disease. The Lancet 397, 1577–1590, doi:10.1016/S0140-6736(20)32205-4 (2021).

2 Duyckaerts, C., Delatour, B. & Potier, M.-C. Classification and basic pathology of Alzheimer disease. Acta neuropathologica 118, 5–36 (2009).

3 Jack Jr, C. R. et al. NIA-AA research framework: toward a biological definition of Alzheimer’s disease. Alzheimer’s & Dementia 14, 535–562 (2018).

4 Mattsson-Carlgren, N. et al. Cerebrospinal fluid biomarkers in autopsy-confirmed Alzheimer disease and frontotemporal lobar degeneration. Neurology 98, e1137–e1150 (2022).

5 Zetterberg, H. & Blennow, K. Moving fluid biomarkers for Alzheimer’s disease from research tools to routine clinical diagnostics. Molecular neurodegeneration 16, 1–7 (2021).

6 Long, J. M. & Holtzman, D. M. Alzheimer disease: an update on pathobiology and treatment strategies. Cell 179, 312–339 (2019).

7 Rayaprolu, S. et al. Systems-based proteomics to resolve the biology of Alzheimer’s disease beyond amyloid and tau. Neuropsychopharmacology 46, 98–115 (2021).

8 Sperling, R. A. et al. Toward defining the preclinical stages of Alzheimer’s disease: Recommendations from the National Institute on Aging-Alzheimer’s Association workgroups on diagnostic guidelines for Alzheimer’s disease. Alzheimer’s & dementia 7, 280–292 (2011).

9 Dubois, B. et al. Preclinical Alzheimer’s disease: definition, natural history, and diagnostic criteria. Alzheimer’s & Dementia 12, 292–323 (2016).

10 Hasin, Y., Seldin, M. & Lusis, A. Multi-omics approaches to disease. Genome biology 18, 1–15 (2017).

11 Seyfried, N. T. et al. A Multi-network Approach Identifies Protein-Specific Co-expression in Asymptomatic and Symptomatic Alzheimer’s Disease. Cell Syst 4, 60–72 e64, doi:10.1016/j.cels.2016.11.006 (2017).

12 Johnson, E. C. B. et al. Deep proteomic network analysis of Alzheimer’s disease brain reveals alterations in RNA binding proteins and RNA splicing associated with disease. Mol Neurodegener 13, 52, doi:10.1186/s13024-018-0282-4 (2018).

13 Johnson, E. C. B. et al. A Consensus Proteomic Analysis of Alzheimer’s Disease Brain and Cerebrospinal Fluid Reveals Early Changes in Energy Metabolism Associated with Microglia and Astrocyte Activation. bioRxiv, 802959, doi:10.1101/802959 (2019).

14 Johnson, E. C. et al. Large-scale proteomic analysis of Alzheimer’s disease brain and cerebrospinal fluid reveals early changes in energy metabolism associated with microglia and astrocyte activation. Nature medicine 26, 769–780 (2020).

15 Blennow, K. & Zetterberg, H. Biomarkers for Alzheimer’s disease: current status and prospects for the future. Journal of internal medicine 284, 643–663 (2018).

16 Higginbotham, L. et al. Integrated proteomics reveals brain-based cerebrospinal fluid biomarkers in asymptomatic and symptomatic Alzheimer’s disease. Science advances 6, eaaz9360 (2020).

17 Picotti, P. & Aebersold, R. Selected reaction monitoring–based proteomics: workflows, potential, pitfalls and future directions. Nature methods 9, 555–566 (2012).

18 Bittner, T. et al. Technical performance of a novel, fully automated electrochemiluminescence immunoassay for the quantitation of beta-amyloid (1-42) in human cerebrospinal fluid. Alzheimers Dement 12, 517–526, doi:10.1016/j.jalz.2015.09.009 (2016).

19 Hansson, O. et al. CSF biomarkers of Alzheimer’s disease concord with amyloid-beta PET and predict clinical progression: A study of fully automated immunoassays in BioFINDER and ADNI cohorts. Alzheimers Dement 14, 1470–1481, doi:10.1016/j.jalz.2018.01.010 (2018).

20 Schindler, S. E. et al. Cerebrospinal fluid biomarkers measured by Elecsys assays compared to amyloid imaging. Alzheimers Dement, doi:10.1016/j.jalz.2018.01.013 (2018).

21 Zhou, M. et al. Targeted mass spectrometry to quantify brain-derived cerebrospinal fluid biomarkers in Alzheimer’s disease. Clinical Proteomics 17, 1–14 (2020).

22 Watson, C. M., Dammer, E.B., and Seyfried, N.T. Emory AD CSF SRM. Synapse, doi:https://doi.org/10.7303/syn34054965 (2022).

23 MacLean, B. et al. Skyline: an open source document editor for creating and analyzing targeted proteomics experiments. Bioinformatics 26, 966–968 (2010).

24 Pino, L. K. et al. The Skyline ecosystem: Informatics for quantitative mass spectrometry proteomics. Mass spectrometry reviews 39, 229–244 (2020).

25 MacLean, B. et al. Effect of collision energy optimization on the measurement of peptides by selected reaction monitoring (SRM) mass spectrometry. Analytical chemistry 82, 10116–10124 (2010).

26 Kulak, N. A., Pichler, G., Paron, I., Nagaraj, N. & Mann, M. Minimal, encapsulated proteomic-sample processing applied to copy-number estimation in eukaryotic cells. Nature methods 11, 319–324 (2014).

27 Simon, R. et al. Total ApoE and ApoE4 isoform assays in an Alzheimer’s disease casecontrol study by targeted mass spectrometry (n= 669): a pilot assay for methioninecontaining proteotypic peptides. Molecular & Cellular Proteomics 11, 1389–1403 (2012).

28 Rezeli, M. et al. Quantification of total apolipoprotein E and its specific isoforms in cerebrospinal fluid and blood in Alzheimer’s disease and other neurodegenerative diseases. EuPA Open Proteomics 8, 137–143 (2015).

29 Geyer, P. E. et al. Plasma Proteome Profiling to detect and avoid sample-related biases in biomarker studies. EMBO molecular medicine 11, e10427 (2019).

30 Beri, J., Rosenblatt, M. M., Strauss, E., Urh, M. & Bereman, M. S. Reagent for evaluating liquid chromatography–tandem mass spectrometry (LC-MS/MS) performance in bottom-up proteomic experiments. Analytical chemistry 87, 11635–11640 (2015).

31 Hatters, D. M., Peters-Libeu, C. A. & Weisgraber, K. H. Apolipoprotein E structure: insights into function. Trends in biochemical sciences 31, 445–454 (2006).

32 Saunders, A. M. et al. Association of apolipoprotein E allele ϵ4 with late-onset familial and sporadic Alzheimer’s disease. Neurology 43, 1467–1467 (1993).

33 Farrer, L. A. et al. Effects of age, sex, and ethnicity on the association between apolipoprotein E genotype and Alzheimer disease. A meta-analysis. APOE and Alzheimer Disease Meta Analysis Consortium. JAMA 278, 1349–1356 (1997).

34 Heffernan, A. L., Chidgey, C., Peng, P., Masters, C. L. & Roberts, B. R. The neurobiology and age-related prevalence of the ε4 allele of apolipoprotein E in Alzheimer’s disease cohorts. Journal of Molecular Neuroscience 60, 316–324 (2016).

35 Minta, K. et al. Quantification of total apolipoprotein E and its isoforms in cerebrospinal fluid from patients with neurodegenerative diseases. Alzheimer’s research & therapy 12, 1–11 (2020).

36 Bussy, A. et al. Effect of apolipoprotein E4 on clinical, neuroimaging, and biomarker measures in noncarrier participants in the Dominantly Inherited Alzheimer Network. Neurobiology of aging 75, 42–50 (2019).

37 El-Lebedy, D., Raslan, H. M. & Mohammed, A. M. Apolipoprotein E gene polymorphism and risk of type 2 diabetes and cardiovascular disease. Cardiovascular diabetology 15, 1–11 (2016).

38 Morris, J. C. et al. APOE predicts amyloid-beta but not tau Alzheimer pathology in cognitively normal aging. Ann Neurol 67, 122–131, doi:10.1002/ana.21843 (2010).

39 Grimmer, T. et al. Progression of cerebral amyloid load is associated with the apolipoprotein E ε4 genotype in Alzheimer’s disease. Biological psychiatry 68, 879–884 (2010).

40 Knopman, D. S. et al. The National Institute on Aging and the Alzheimer’s Association research framework for Alzheimer’s disease: perspectives from the research roundtable. Alzheimer’s & Dementia 14, 563–575 (2018).

41 Wingo, A. P. et al. Large-scale proteomic analysis of human brain identifies proteins associated with cognitive trajectory in advanced age. Nature communications 10, 1–14 (2019).

42 Dammer, E. B. et al. Multi-Platform Proteomic Analysis of Alzheimer’s Disease Cerebrospinal Fluid and Plasma Reveals Network Biomarkers Associated with Proteostasis and the Matrisome. bioRxiv (2022).

43 Bertens, D., Knol, D. L., Scheltens, P. & Visser, P. J. Temporal evolution of biomarkers and cognitive markers in the asymptomatic, MCI, and dementia stage of Alzheimer’s disease. Alzheimer’s & Dementia 11, 511–522, doi:https://doi.org/10.1016/j.jalz.2014.05.1754 (2015).

44 Shokouhi, S. et al. Reference tissue normalization in longitudinal 18F-florbetapir positron emission tomography of late mild cognitive impairment. Alzheimer’s Research & Therapy 8, 2, doi:10.1186/s13195-016-0172-3 (2016).

45 Sathe, G. et al. Quantitative proteomic profiling of cerebrospinal fluid to identify candidate biomarkers for Alzheimer’s disease. PROTEOMICS–Clinical Applications 13, 1800105 (2019).

46 Bader, J. M. et al. Proteome profiling in cerebrospinal fluid reveals novel biomarkers of Alzheimer’s disease. Molecular systems biology 16, e9356 (2020).

47 Libiger, O. et al. Longitudinal CSF proteomics identifies NPTX2 as a prognostic biomarker of Alzheimer’s disease. Alzheimer’s & Dementia 17, 1976–1987 (2021).

48 Johnson, E. C. et al. Large-scale deep multi-layer analysis of Alzheimer’s disease brain reveals strong proteomic disease-related changes not observed at the RNA level. Nature neuroscience 25, 213–225 (2022).

